# Effectiveness of BNT162b2 and mRNA-1273 COVID-19 boosters against SARS-CoV-2 Omicron (B.1.1.529) infection in Qatar

**DOI:** 10.1101/2022.01.18.22269452

**Authors:** Laith J. Abu-Raddad, Hiam Chemaitelly, Houssein H. Ayoub, Sawsan AlMukdad, Patrick Tang, Mohammad R. Hasan, Peter Coyle, Hadi M. Yassine, Hebah A. Al-Khatib, Maria K. Smatti, Zaina Al-Kanaani, Einas Al-Kuwari, Andrew Jeremijenko, Anvar Hassan Kaleeckal, Ali Nizar Latif, Riyazuddin Mohammad Shaik, Hanan F. Abdul-Rahim, Gheyath K. Nasrallah, Mohamed Ghaith Al-Kuwari, Adeel A. Butt, Hamad Eid Al-Romaihi, Mohamed H. Al-Thani, Abdullatif Al-Khal, Roberto Bertollini

## Abstract

**BACKGROUND:** Waning of COVID-19 vaccine protection and emergence of SARS-CoV-2 Omicron (B.1.1.529) variant have expedited efforts to scale up booster vaccination. This study compared protection afforded by booster doses of the BNT162b2 (Pfizer-BioNTech) and mRNA-1273 (Moderna) vaccines, compared to the primary series of only two doses in Qatar, during a large, rapidly growing Omicron wave.

**METHODS:** In a population of 2,232,224 vaccinated persons with at least two doses, two matched, retrospective cohort studies were implemented to investigate effectiveness of booster vaccination against symptomatic SARS-CoV-2 infection and against COVID-19 hospitalization and death, up to January 9, 2022. Association of booster status with infection was estimated using Cox proportional-hazards regression models.

**RESULTS:** For BNT162b2, cumulative symptomatic infection incidence was 2.9% (95% CI: 2.8-3.1%) in the booster-dose cohort and 5.5% (95% CI: 5.3-5.7%) in the primary-series cohort, after 49 days of follow-up. Adjusted hazard ratio for symptomatic infection was 0.50 (95% CI: 0.47-0.53). Booster effectiveness relative to primary series was 50.1% (95% CI: 47.3-52.8%). For mRNA-1273, cumulative symptomatic infection incidence was 1.9% (95% CI: 1.7-2.2%) in the booster-dose cohort and 3.5% (95% CI: 3.2-3.9%) in the primary-series cohort, after 35 days of follow-up. The adjusted hazard ratio for symptomatic infection was 0.49 (95% CI: 0.43-0.57). Booster effectiveness relative to primary series was 50.8% (95% CI: 43.4-57.3%). There were fewer cases of severe COVID-19 in booster-dose cohorts than in primary-series cohorts, but cases of severe COVID-19 were rare in all cohorts.

**CONCLUSIONS:** mRNA booster vaccination is associated with modest effectiveness against symptomatic infection with Omicron. The development of a new generation of vaccines targeting a broad range of variants may be warranted.

## Introduction

Waning of coronavirus disease 2019 (COVID-19) vaccine protection^1-6^ and emergence of the immune-evasive severe acute respiratory syndrome coronavirus 2 (SARS-CoV-2) Omicron^7^ (B.1.1.529) variant of concern^8-10^ have stimulated efforts to scale-up COVID-19 booster vaccination. Qatar launched its third dose/booster vaccination program in mid-September 2021, using both the BNT162b2^11^ (Pfizer-BioNTech) and mRNA-1273^12^ (Moderna) mRNA vaccines, the same two vaccines that have been used since the launch of the COVID-19 immunization program in this country.^13-15^

Initially, booster eligibility was restricted to the elderly, immunocompromised persons, and persons with severe or multiple chronic conditions, but then expanded by age group to the rest of the population. Eligibility was first restricted to persons who had completed the primary series of two doses at least 8 months earlier, but subsequently this interval was shortened to 6 months. The same vaccine was used in both primary series and booster vaccinations for the majority of the population, but for mRNA-1273, the booster dose was half the dose used in the primary series. As booster vaccination was being scaled up, the Omicron variant was introduced into the population, leading to the largest SARS-CoV-2 epidemic wave that the country has experienced since the start of the pandemic.

Leveraging the integrated national COVID-19 datasets, effectiveness of booster vaccination against SARS-CoV-2 symptomatic infection and against COVID-19 hospitalization and death, relative to that of the primary series of two doses, was investigated in the national cohort of vaccinated individuals in Qatar, during the rapidly growing Omicron wave.^10^

## Methods

### Data sources and study design

The study analyzed the national, federated databases for COVID-19 vaccination, laboratory testing, hospitalization, and death, retrieved from the integrated nationwide digital-health information platform. Databases include all SARS-CoV-2-related data and associated demographic information, with no missing information, since pandemic onset. These include all polymerase chain reaction (PCR) tests, vaccination records, COVID-19 hospitalizations, infection severity and mortality classifications per World Health Organization (WHO) guidelines,^16,17^ in addition to sex, age, and nationality information retrieved from the national registry. Further description of these national databases can be found in previous studies.^1,13,14,18-24^

Effectiveness of booster vaccination relative to the primary series was estimated for both the BNT162b2 and mRNA-1273 vaccines using a matched, retrospective cohort study design that emulated a target trial.^25,26^ The study compared incidence of SARS-CoV-2 symptomatic breakthrough infection in the cohort of individuals who completed >7 days after the booster dose (booster-dose cohort) to incidence in the cohort of individuals who have not yet received their booster dose (primary-series cohort). The 7-day cutoff between administration of the booster and start of follow-up was informed by earlier studies^26-29^ to ensure sufficient time for build-up of booster protection. A 14-day cutoff was also investigated in a sensitivity analysis.

For BNT162b2, all individuals with at least two doses between January 5, 2021 (date of first second-dose BNT162b2 vaccination in Qatar) and January 9, 2022 (end of study) were eligible for inclusion in the study, provided that they had no record of a prior PCR-confirmed infection before the start of follow-up. The same applied to mRNA-1273, but the corresponding dates were January 24, 2021 and January 9, 2022, respectively.

Individuals in the booster-dose cohort were exact-matched in a 1:1 ratio by sex, 10-year age group, and nationality to individuals in the primary-series cohort, to control for known differences in the risk of exposure to SARS-CoV-2 infection in Qatar.^18,21,30-32^ Matching by these factors was shown in an earlier study of similar design to provide adequate control of bias arising from differences in infection exposure in Qatar.^3^ Individuals were also exact-matched by the calendar week of second vaccine dose to control for time since vaccination and waning of vaccine immunity over time.^1,3,6,15^

Matching was performed through an iterative process that ensured that each control in the primary-series cohort is alive, infection-free, and has not received the third dose at the start of follow-up, which was defined, for each matched pair, at the 8^th^ day after the individual in the booster-dose cohort received the booster dose. Controls in the primary-series cohort who received the booster dose at a future date were eligible for recruitment into the booster-dose cohort, provided that they were alive and infection-free at the start of follow-up. Accordingly, some individuals contributed follow-up time both as persons with only a primary series and as persons with a booster, but at different times.

Both members of each matched pair were censored at the event of the control receiving the booster dose, to ensure exchangeability.^26^ Accordingly, individuals were followed up until the first of one of the following events: a documented SARS-CoV-2 infection (defined as the first PCR-positive test after the start of follow-up regardless of presence of symptoms or reason for PCR testing), or booster-dose vaccination of the control (with matched pair censoring), death, or end of study censoring (January 9, 2022).

Investigated primary outcome was symptomatic infection defined as a PCR-positive nasopharyngeal swab conducted because of clinical suspicion due to presence of symptoms compatible with a respiratory tract infection. Investigated secondary outcome was any severe,^16^ critical,^16^ or fatal^17^ COVID-19. Classification of COVID-19 case severity (acute-care hospitalizations),^16^ criticality (ICU hospitalizations),^16^ and fatality^17^ followed WHO guidelines, and assessments were made by trained medical personnel using individual chart reviews. Details of the COVID-19 severity, criticality, and fatality classification are found in Section 1 in Supplementary Appendix.

Each person hospitalized for COVID-19 underwent an infection severity assessment every three days until discharge or death. We classified individuals who progressed to severe, critical, or fatal COVID-19 between the time of the PCR-positive test and the end of the study based on their worst outcome, starting with death,^17^ followed by critical disease,^16^ and then severe disease.^16^

### Laboratory methods

Details of laboratory methods for real-time reverse-transcription PCR (RT-qPCR) testing are found in Section 2 in Supplementary Appendix.

### Statistical analysis

Frequency distributions and measures of central tendency were used to describe the full and matched cohorts. Group comparison was performed using standardized mean differences (SMDs), with an SMD <0.1 indicating adequate matching.^33^ Cumulative incidence of symptomatic infection was defined as the proportion of individuals at risk whose primary endpoint was a documented symptomatic infection during follow-up, and was estimated in each cohort using the Kaplan–Meier estimator method.^34^ Equality of failure functions was assessed using the log-rank test. Incidence rate of symptomatic infection in each cohort, which was defined as the number of identified symptomatic infections divided by the number of person-weeks contributed by all individuals in the cohort, was estimated, along with its 95% confidence interval (CI), using a Poisson log-likelihood regression model with the STATA 17.0^35^ *stptime* command.

The hazard ratio comparing incidence of symptomatic infection in both cohorts and corresponding 95% CI were calculated using Cox regression adjusted for the matching factors with the STATA 17.0^35^ *stcox* command. Shoenfeld residuals and log-log plots for the survival curves were used to test the proportional-hazards assumption and to investigate its adequacy. 95% CIs were not adjusted for multiplicity. Interactions were not considered. Vaccine effectiveness of booster vaccination relative to the primary series was estimated using the equation: Vaccine effectiveness = 1−adjusted hazrad ratio.

In addition to estimating the booster-vaccine effectiveness against symptomatic infection with Omicron, an additional sub-analysis was conducted to estimate effectiveness against symptomatic infection with Delta (B.1.617.2).^7^ Here, the end of the study period was December 1, 2021, that is at first detection of Omicron in Qatar.^10,36^ During this specific follow-up period, Qatar was experiencing a Delta-dominated low-incidence phase.^1,13,14,23,36-38^ This analysis was only possible for the BNT162b2 vaccine as the time of follow-up was limited for the mRNA-1273-vaccine cohorts.

Since optimal effectiveness of the booster dose may require more than 7 days to develop, the main analyses for the BNT162b2 and mRNA-1273 vaccines were repeated, but with the start of follow-up on the 15^th^ day after receiving the booster dose, instead of the 8^th^ day.

Booster-vaccine effectiveness was also calculated against any documented infection regardless of presence of symptoms or reason for PCR testing.

Statistical analyses were conducted in STATA/SE version 17.0.^35^

### Oversight

Hamad Medical Corporation and Weill Cornell Medicine-Qatar Institutional Review Boards approved this retrospective study with waiver of informed consent, since analysis was done on routinely collected data. The study was reported following the Strengthening the Reporting of Observational Studies in Epidemiology (STROBE) guidelines. The STROBE checklist can be found in Table S1 of the Supplementary Appendix.

## Results

### Study population for BNT162b2 vaccine

Between January 5, 2021 and January 9, 2022, 1,294,138 individuals received at least two doses of BNT162b2 and 230,526 received a third, BNT162b2 booster dose. The median date at the first dose was May 3, 2021, May 24, 2021 at the second dose, and December 6, 2021 at the third dose. The median time elapsed between the first and second doses was 21 days (interquartile range (IQR), 21-22 days). The median time elapsed between the second and booster doses was 249 days (IQR, 234-267 days).

As of January 9, 2022, 88,729 and 13,948 breakthrough infections were recorded among those who received either only two or three BNT162b2 doses, respectively. Among infections after the second dose, but before the booster dose, 210 progressed to severe, 18 to critical, and 19 to fatal COVID-19. Of the infections after the booster dose, 12 progressed to severe, but none to critical or fatal COVID-19.

Figure 1 shows the population selection process for the BNT162b2-booster study. Table 1 shows baseline characteristics of the full and matched cohorts. Median age in the matched cohorts was 43 years (IQR, 35-53). The matched cohorts were balanced on the matching factors.

**Table 1.**
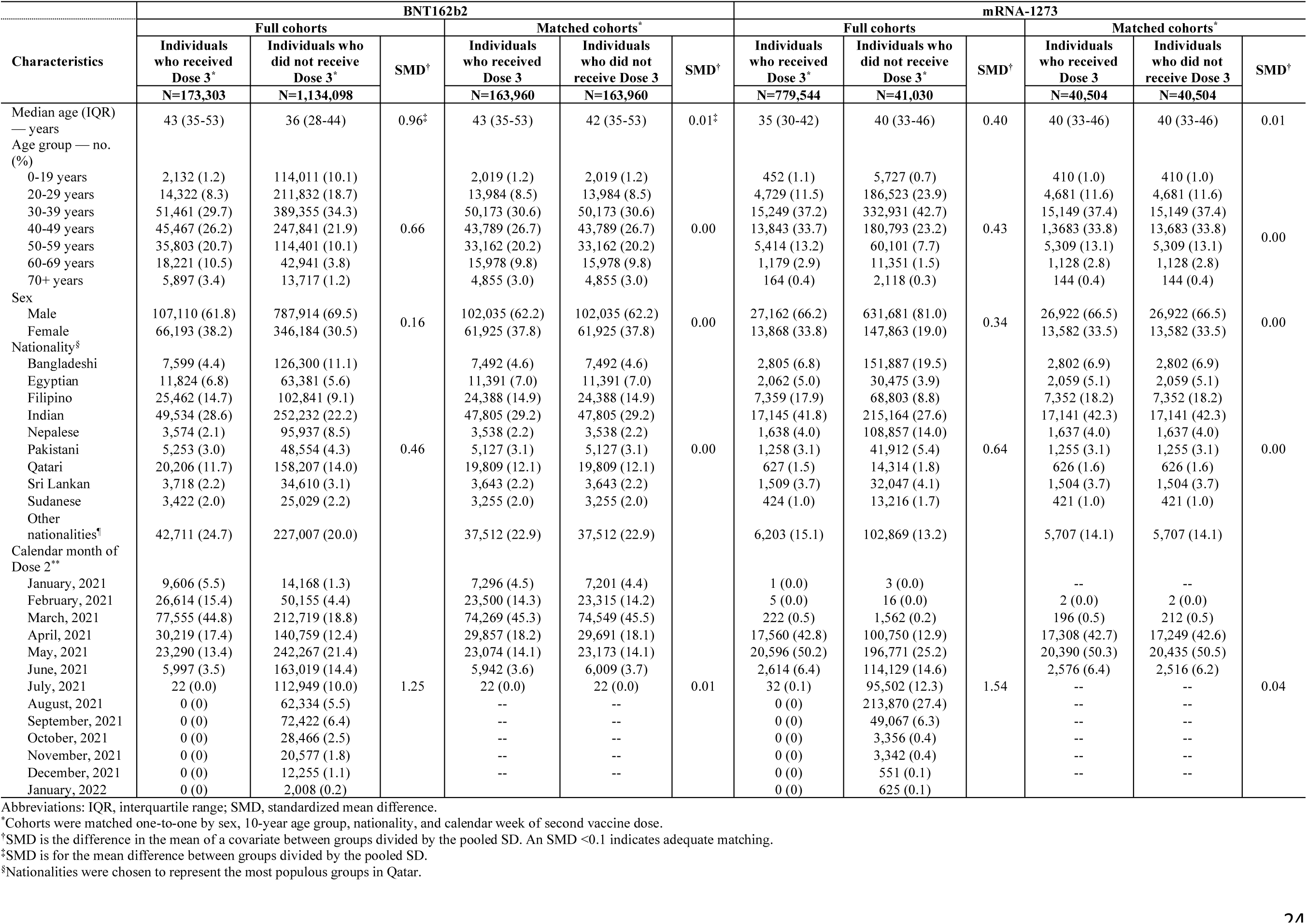
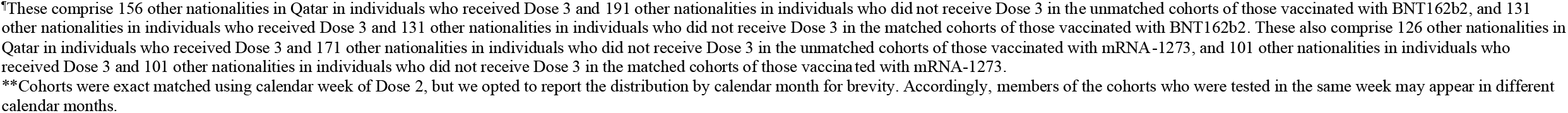
Baseline characteristics of the full and matched primary-series and booster-dose cohorts for the BNT162b2 and mRNA-1273 vaccines.

| Characteristics | BNT162b2 |  |  |  |  |  | mRNA-1273 |  |  |  |  |  |
| --- | --- | --- | --- | --- | --- | --- | --- | --- | --- | --- | --- | --- |
|  | Full cohorts |  |  | Matched cohorts* |  |  | Full cohorts |  |  | Matched cohorts* |  |  |
|  | Individuals who received Dose 3* | Individuals who did not receive Dose 3* | SMD† | Individuals who received Dose 3 | Individuals who did not receive Dose 3 | SMD† | Individuals who received Dose 3* | Individuals who did not receive Dose 3* | SMD† | Individuals who received Dose 3 | Individuals who did not receive Dose 3 | SMD† |
|  | N=173,303 | N=1,134,098 |  | N=163,960 | N=163,960 |  | N=779,544 | N=41,030 |  | N=40,504 | N=40,504 |  |
| Median age (IQR) — years | 43 (35-53) | 36 (28-44) | 0.96‡ | 43 (35-53) | 42 (35-53) | 0.01‡ | 35 (30-42) | 40 (33-46) | 0.40 | 40 (33-46) | 40 (33-46) | 0.01 |
| Age group — no. (%) |  |  |  |  |  |  |  |  |  |  |  |  |
| 0-19 years | 2,132 (1.2) | 114,011 (10.1) |  | 2,019 (1.2) | 2,019 (1.2) |  | 452 (1.1) | 5,727 (0.7) |  | 410 (1.0) | 410 (1.0) |  |
| 20-29 years | 14,322 (8.3) | 211,832 (18.7) |  | 13,984 (8.5) | 13,984 (8.5) |  | 4,729 (11.5) | 186,523 (23.9) |  | 4,681 (11.6) | 4,681 (11.6) |  |
| 30-39 years | 51,461 (29.7) | 389,355 (34.3) |  | 50,173 (30.6) | 50,173 (30.6) |  | 15,249 (37.2) | 332,931 (42.7) |  | 15,149 (37.4) | 15,149 (37.4) |  |
| 40-49 years | 45,467 (26.2) | 247,841 (21.9) | 0.66 | 43,789 (26.7) | 43,789 (26.7) | 0.00 | 13,843 (33.7) | 180,793 (23.2) | 0.43 | 1,3683 (33.8) | 13,683 (33.8) | 0.00 |
| 50-59 years | 35,803 (20.7) | 114,401 (10.1) |  | 33,162 (20.2) | 33,162 (20.2) |  | 5,414 (13.2) | 60,101 (7.7) |  | 5,309 (13.1) | 5,309 (13.1) |  |
| 60-69 years | 18,221 (10.5) | 42,941 (3.8) |  | 15,978 (9.8) | 15,978 (9.8) |  | 1,179 (2.9) | 11,351 (1.5) |  | 1,128 (2.8) | 1,128 (2.8) |  |
| 70+ years | 5,897 (3.4) | 13,717 (1.2) |  | 4,855 (3.0) | 4,855 (3.0) |  | 164 (0.4) | 2,118 (0.3) |  | 144 (0.4) | 144 (0.4) |  |
| Sex |  |  |  |  |  |  |  |  |  |  |  |  |
| Male | 107,110 (61.8) | 787,914 (69.5) | 0.16 | 102,035 (62.2) | 102,035 (62.2) | 0.00 | 27,162 (66.2) | 631,681 (81.0) | 0.34 | 26,922 (66.5) | 26,922 (66.5) | 0.00 |
| Female | 66,193 (38.2) | 346,184 (30.5) |  | 61,925 (37.8) | 61,925 (37.8) |  | 13,868 (33.8) | 147,863 (19.0) |  | 13,582 (33.5) | 13,582 (33.5) |  |
| Nationality§ |  |  |  |  |  |  |  |  |  |  |  |  |
| Bangladeshi | 7,599 (4.4) | 126,300 (11.1) |  | 7,492 (4.6) | 7,492 (4.6) |  | 2,805 (6.8) | 151,887 (19.5) |  | 2,802 (6.9) | 2,802 (6.9) |  |
| Egyptian | 11,824 (6.8) | 63,381 (5.6) |  | 11,391 (7.0) | 11,391 (7.0) |  | 2,062 (5.0) | 30,475 (3.9) |  | 2,059 (5.1) | 2,059 (5.1) |  |
| Filipino | 25,462 (14.7) | 102,841 (9.1) |  | 24,388 (14.9) | 24,388 (14.9) |  | 7,359 (17.9) | 68,803 (8.8) |  | 7,352 (18.2) | 7,352 (18.2) |  |
| Indian | 49,534 (28.6) | 252,232 (22.2) |  | 47,805 (29.2) | 47,805 (29.2) |  | 17,145 (41.8) | 215,164 (27.6) |  | 17,141 (42.3) | 17,141 (42.3) |  |
| Nepalese | 3,574 (2.1) | 95,937 (8.5) |  | 3,538 (2.2) | 3,538 (2.2) |  | 1,638 (4.0) | 108,857 (14.0) |  | 1,637 (4.0) | 1,637 (4.0) |  |
| Pakistani | 5,253 (3.0) | 48,554 (4.3) | 0.46 | 5,127 (3.1) | 5,127 (3.1) | 0.00 | 1,258 (3.1) | 41,912 (5.4) | 0.64 | 1,255 (3.1) | 1,255 (3.1) | 0.00 |
| Qatari | 20,206 (11.7) | 158,207 (14.0) |  | 19,809 (12.1) | 19,809 (12.1) |  | 627 (1.5) | 14,314 (1.8) |  | 626 (1.6) | 626 (1.6) |  |
| Sri Lankan | 3,718 (2.2) | 34,610 (3.1) |  | 3,643 (2.2) | 3,643 (2.2) |  | 1,509 (3.7) | 32,047 (4.1) |  | 1,504 (3.7) | 1,504 (3.7) |  |
| Sudanese | 3,422 (2.0) | 25,029 (2.2) |  | 3,255 (2.0) | 3,255 (2.0) |  | 424 (1.0) | 13,216 (1.7) |  | 421 (1.0) | 421 (1.0) |  |
| Other nationalities¶ | 42,711 (24.7) | 227,007 (20.0) |  | 37,512 (22.9) | 37,512 (22.9) |  | 6,203 (15.1) | 102,869 (13.2) |  | 5,707 (14.1) | 5,707 (14.1) |  |
| Calendar month of Dose 2** |  |  |  |  |  |  |  |  |  |  |  |  |
| January, 2021 | 9,606 (5.5) | 14,168 (1.3) |  | 7,296 (4.5) | 7,201 (4.4) |  | 1 (0.0) | 3 (0.0) |  | -- | -- |  |
| February, 2021 | 26,614 (15.4) | 50,155 (4.4) |  | 23,500 (14.3) | 23,315 (14.2) |  | 5 (0.0) | 16 (0.0) |  | 2 (0.0) | 2 (0.0) |  |
| March, 2021 | 77,555 (44.8) | 212,719 (18.8) |  | 74,269 (45.3) | 74,549 (45.5) |  | 222 (0.5) | 1,562 (0.2) |  | 196 (0.5) | 212 (0.5) |  |
| April, 2021 | 30,219 (17.4) | 140,759 (12.4) |  | 29,857 (18.2) | 29,691 (18.1) |  | 17,560 (42.8) | 100,750 (12.9) |  | 17,308 (42.7) | 17,249 (42.6) |  |
| May, 2021 | 23,290 (13.4) | 242,267 (21.4) |  | 23,074 (14.1) | 23,173 (14.1) |  | 20,596 (50.2) | 196,771 (25.2) |  | 20,390 (50.3) | 20,435 (50.5) |  |
| June, 2021 | 5,997 (3.5) | 163,019 (14.4) |  | 5,942 (3.6) | 6,009 (3.7) |  | 2,614 (6.4) | 114,129 (14.6) |  | 2,576 (6.4) | 2,516 (6.2) |  |
| July, 2021 | 22 (0.0) | 112,949 (10.0) | 1.25 | 22 (0.0) | 22 (0.0) | 0.01 | 32 (0.1) | 95,502 (12.3) | 1.54 | -- | -- | 0.04 |
| August, 2021 | 0 (0) | 62,334 (5.5) |  | -- | -- |  | 0 (0) | 213,870 (27.4) |  | -- | -- |  |
| September, 2021 | 0 (0) | 72,422 (6.4) |  | -- | -- |  | 0 (0) | 49,067 (6.3) |  | -- | -- |  |
| October, 2021 | 0 (0) | 28,466 (2.5) |  | -- | -- |  | 0 (0) | 3,356 (0.4) |  | -- | -- |  |
| November, 2021 | 0 (0) | 20,577 (1.8) |  | -- | -- |  | 0 (0) | 3,342 (0.4) |  | -- | -- |  |
| December, 2021 | 0 (0) | 12,255 (1.1) |  | -- | -- |  | 0 (0) | 551 (0.1) |  | -- | -- |  |
| January, 2022 | 0 (0) | 2,008 (0.2) |  | -- | -- |  | 0 (0) | 625 (0.1) |  | -- | -- |  |

Abbreviations: IQR, interquartile range; SMD, standardized mean difference.
\*Cohorts were matched one-to-one by sex, 10-year age group, nationality, and calendar week of second vaccine dose.
†SMD is the difference in the mean of a covariate between groups divided by the pooled SD. An SMD <0.1 indicates adequate matching.
‡SMD is for the mean difference between groups divided by the pooled SD.
§Nationalities were chosen to represent the most populous groups in Qatar.
\*These comprise 156 other nationalities in Qatar in individuals who received Dose 3 and 191 other nationalities in individuals who did not receive Dose 3 in the unmatched cohorts of those vaccinated with BNT162b2, and 131 other nationalities in individuals who received Dose 3 and 131 other nationalities in individuals who did not receive Dose 3 in the matched cohorts of those vaccinated with BNT162b2. These also comprise 126 other nationalities in Qatar in individuals who received Dose 3 and 171 other nationalities in individuals who did not receive Dose 3 in the unmatched cohorts of those vaccinated with mRNA-1273, and 101 other nationalities in individuals who received Dose 3 and 101 other nationalities in individuals who did not receive Dose 3 in the matched cohorts of those vaccinated with mRNA-1273.
\*\*Cohorts were exact matched using calendar week of Dose 2, but we opted to report the distribution by calendar month for brevity. Accordingly, members of the cohorts who were tested in the same week may appear in different calendar months.

**Figure 1.**
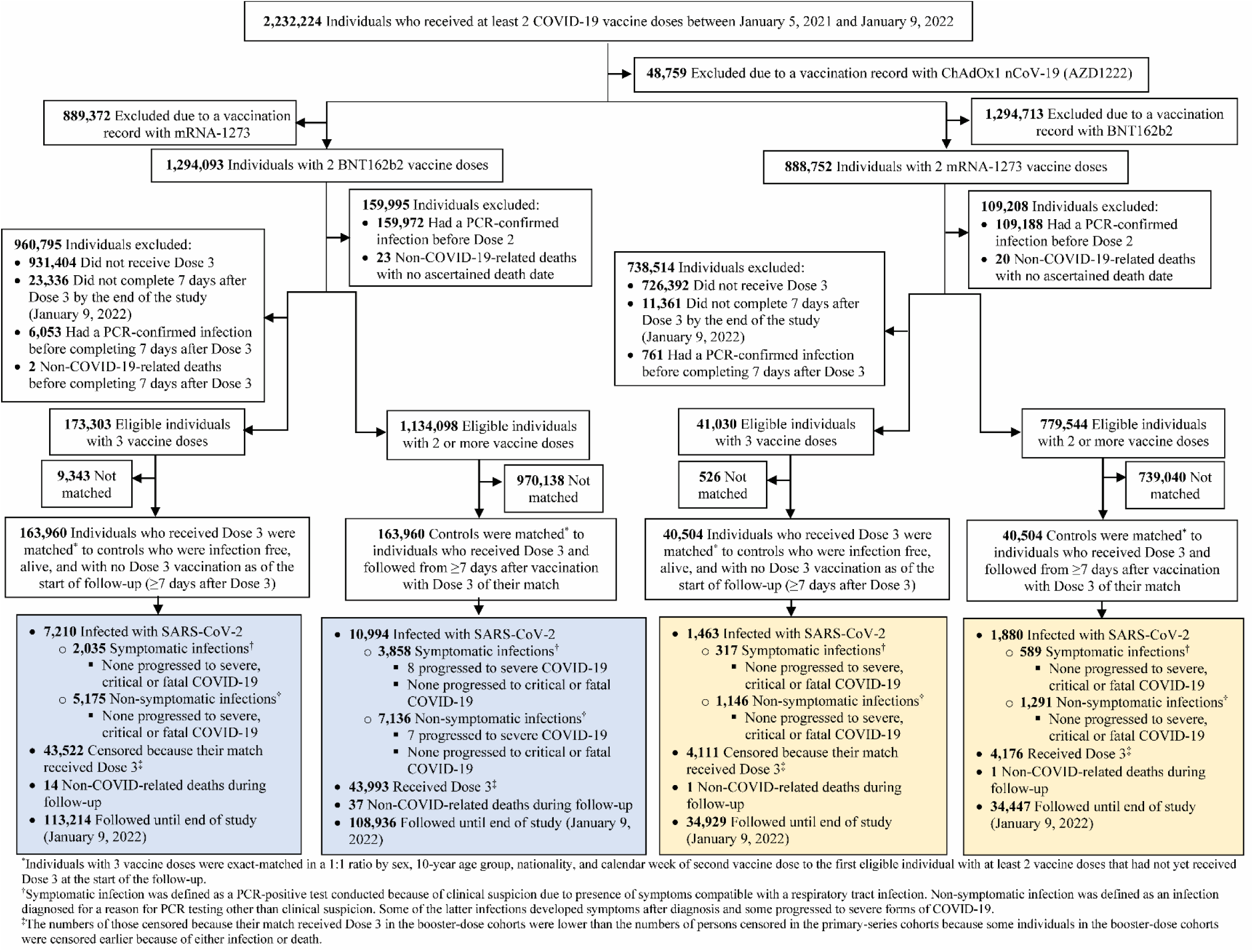
Cohort selection in studying effectiveness of BNT162b2 and mRNA-1273 booster vaccination.

### Effectiveness of BNT162b2 booster against Omicron

The median time of follow-up was 21 days (IQR, 8-36 days) for the booster-dose and primary-series BNT162b2 cohorts. A total of 7,210 infections, 2,035 symptomatic and 5,175 non-symptomatic at diagnosis, were recorded in the booster-dose cohort 8 days or more after receiving the booster dose (Figure 1). None of these infections progressed to severe, critical, or fatal COVID-19 as of end of this study. Although COVID-19 hospitalizations occurred among those who received a booster dose (note section above), all of these cases occurred due to infections within the first 7 days after the booster dose, after censoring for those being followed, or among those not part of the matched cohort—none of these cases were due to infections during time of follow-up of the matched cohort.

A total of 10,994 infections, 3,858 symptomatic and 7,136 non-symptomatic, were recorded in the primary-series cohort. Of these infections, 15 progressed to severe COVID-19, but none to critical or fatal COVID-19.

Cumulative incidence of symptomatic infection was estimated at 2.9% (95% CI: 2.8-3.1%) for the booster-dose cohort and at 5.5% (95% CI: 5.3-5.7%) for the primary-series cohort, 49 days after the start of follow-up (Figure 2). Infection incidence was overwhelmingly predominated by Omicron. The median start date of follow-up was December 9, 2021, after introduction of Omicron in Qatar, but a few days before onset of the large Omicron-wave exponential-growth phase on December 19, 2021.^10,36^ Of all PCR-documented infections between December 1, 2021 and January 9, 2022 (end of study), that is the duration over which most time of follow-up occurred, 97.5% of infections occurred after onset of the Omicron-wave exponential-growth phase. Of 98 sequenced random specimens that were collected between December 19 and December 31, 2021, 86 (88%) were confirmed as Omicron infections and 12 (12%) as Delta infections.^36^

**Figure 2.**
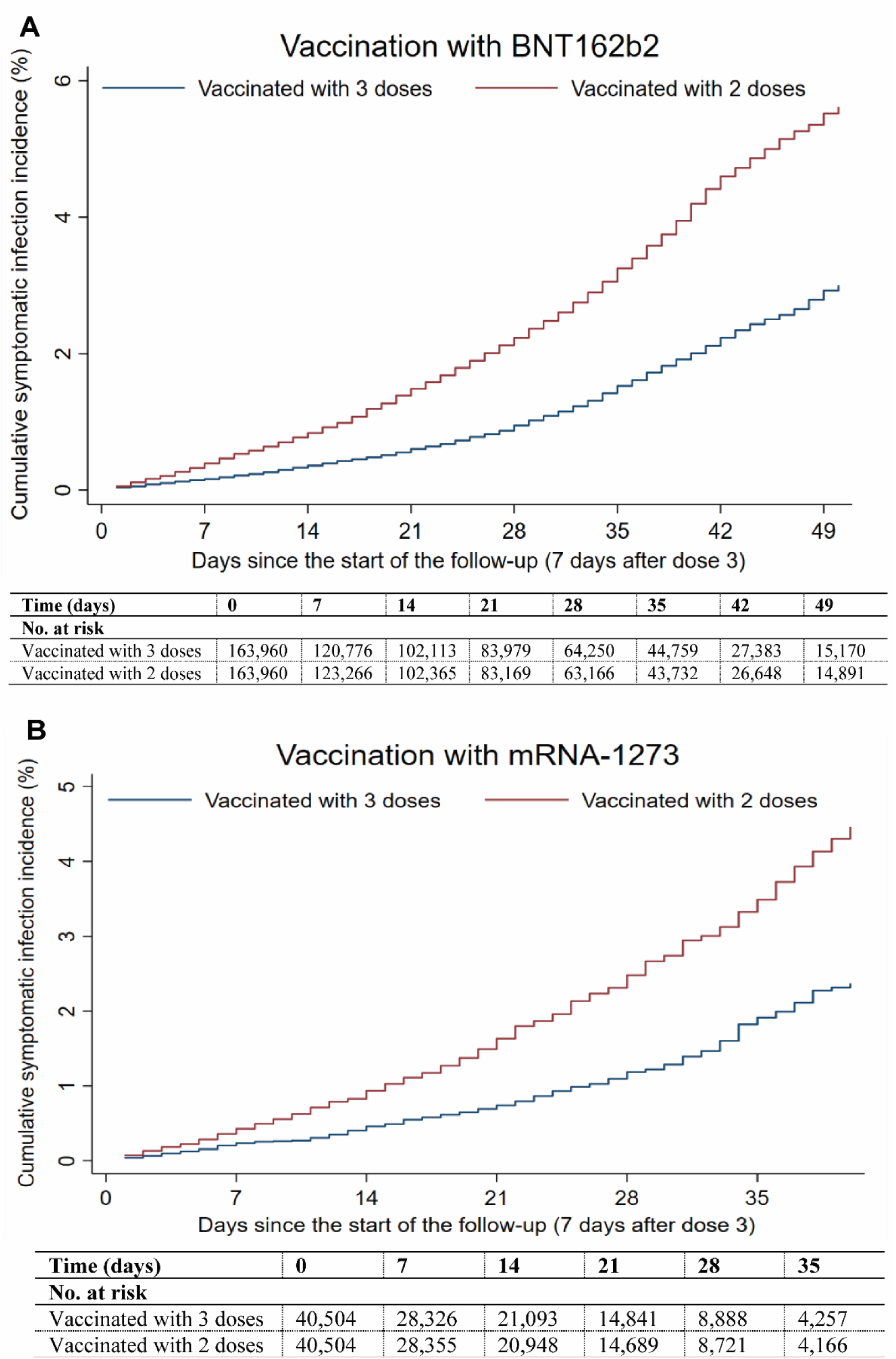
Cumulative incidence of symptomatic infection in the matched booster-dose and primary-series cohorts for the BNT162b2 (A) and mRNA-1273 (B) vaccines.

Symptomatic infection incidence rate was estimated at 35.1 (95% CI: 33.6-36.6) per 10,000 person-weeks in the booster-dose cohort and at 67.2 (95% CI: 65.1-69.3) per 10,000 person-weeks in the primary-series cohort (Table 2). The hazard ratio for symptomatic infection, adjusted for sex, 10-year age group, nationality group, and calendar week of dose 2, was estimated at 0.50 (95% CI: 0.47-0.53). Effectiveness of BNT162b2 booster dose relative to the primary series of only two doses was estimated at 50.1% (95% CI: 47.3-52.8%).

**Table 2.**
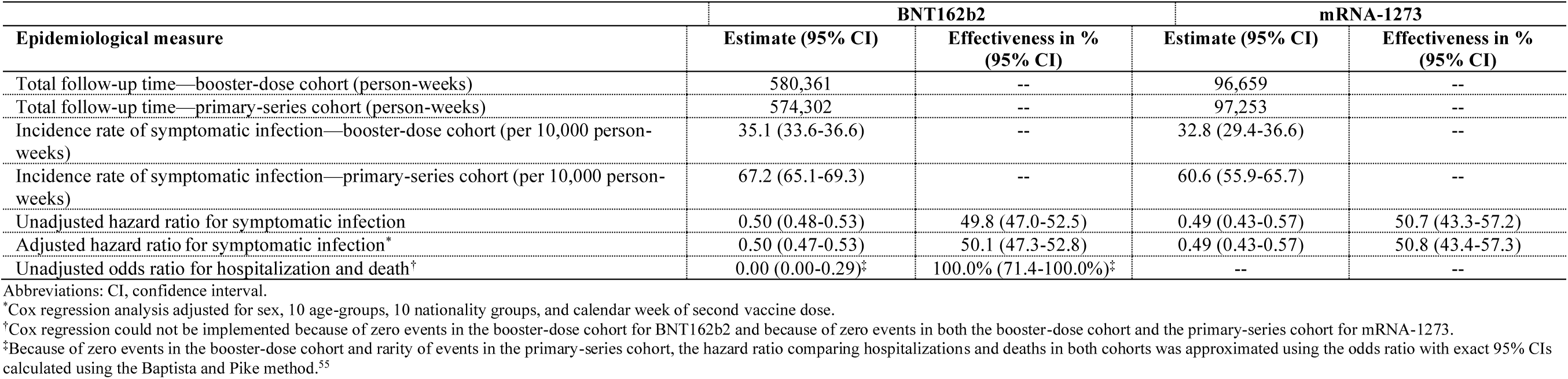
Effectiveness of BNT162b2 and mRNA-1273 booster vaccination.

The hazard ratio for any severe, critical, or fatal COVID-19 was estimated at 0.00 (95% CI: 0.00-0.29). Effectiveness of the BNT162b2 booster dose against any severe, critical, or fatal COVID-19, relative to the primary series, was estimated at 100.0% (95% CI: 71.4-100.0%).

### Effectiveness of BNT162b2 booster against Delta

In the Delta-variant analysis, that is with the end of study on December 1, 2021, cumulative incidence of symptomatic infection was estimated at 0.05% (95% CI: 0.02-0.1%) for the BNT162b2 booster-dose cohort and at 0.2% (95% CI: 0.1-0.3%) for the primary-series cohort, 49 days after the start of follow-up (Figure 3). Symptomatic infection incidence rate was estimated at 0.6 (95% CI: 0.3-1.2) per 10,000 person-weeks in the booster-dose cohort and at 3.6 (95% CI: 2.6-4.8) per 10,000 person-weeks in the primary-series cohort (Table 3). The adjusted hazard ratio for symptomatic infection was estimated at 0.14 (95% CI: 0.06-0.33). Effectiveness of BNT162b2 booster dose relative to the primary series was estimated at 86.1% (95% CI: 67.3-94.1%).

**Table 3.**
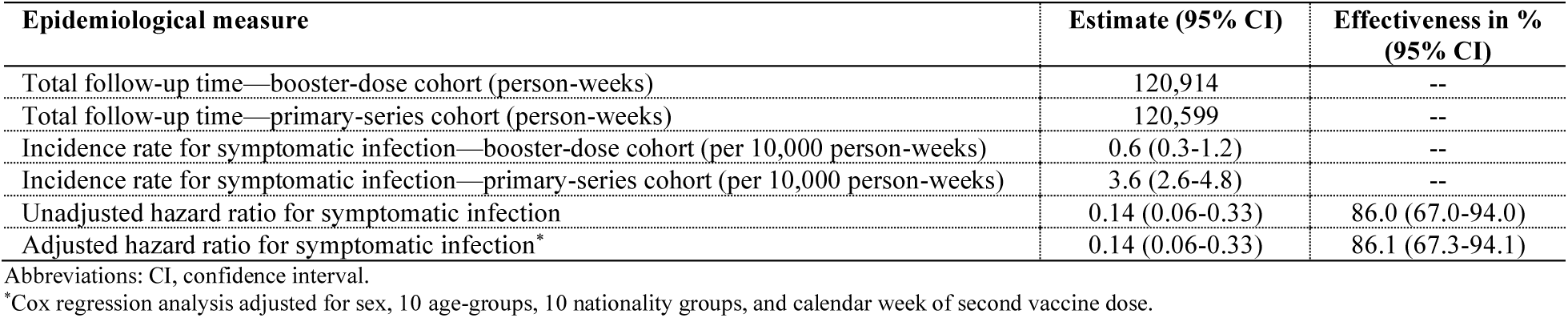
Effectiveness of BNT162b2 booster vaccination during the Delta-predominated low-incidence phase.

| Epidemiological measure | Estimate (95% CI) | Effectiveness in %<br>(95% CI) |
| --- | --- | --- |
| Total follow-up time—booster-dose cohort (person-weeks) | 120,914 | -- |
| Total follow-up time—primary-series cohort (person-weeks) | 120,599 | -- |
| Incidence rate for symptomatic infection—booster-dose cohort (per 10,000 person-weeks) | 0.6 (0.3-1.2) | -- |
| Incidence rate for symptomatic infection—primary-series cohort (per 10,000 person-weeks) | 3.6 (2.6-4.8) | -- |
| Unadjusted hazard ratio for symptomatic infection | 0.14 (0.06-0.33) | 86.0 (67.0-94.0) |
| Adjusted hazard ratio for symptomatic infection<sup>a</sup> | 0.14 (0.06-0.33) | 86.1 (67.3-94.1) |

Abbreviations: CI, confidence interval.
<sup>a</sup>Cox regression analysis adjusted for sex, 10 age-groups, 10 nationality groups, and calendar week of second vaccine dose.

**Figure 3.**
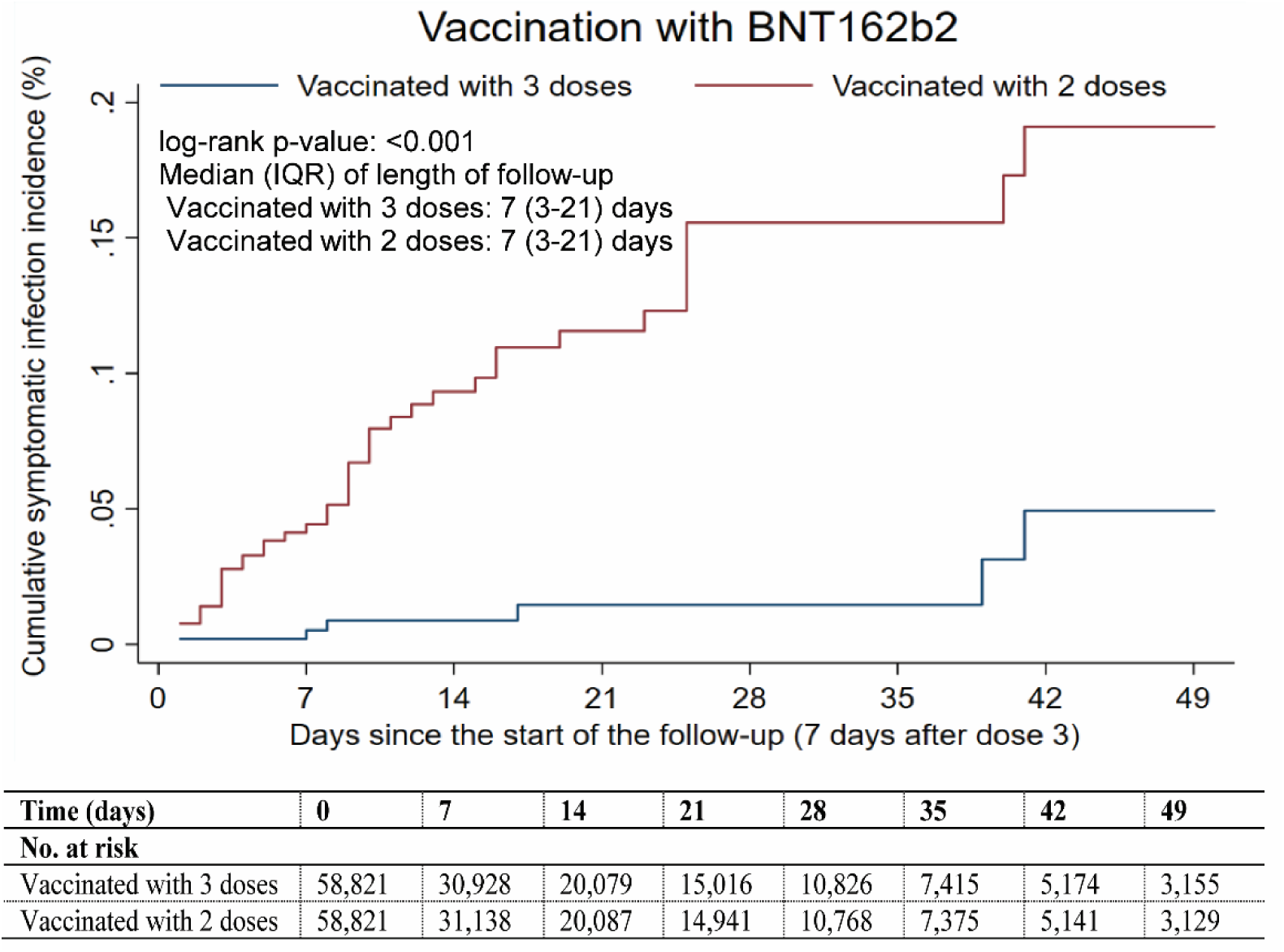
Cumulative incidence of symptomatic infection in the matched booster-dose and primary-series cohorts for the BNT162b2 vaccine during the Delta-predominated low-incidence phase.

### Study population for mRNA-1273 vaccine

Between January 24, 2021 and January 9, 2022, 889,022 individuals received at least two doses of mRNA-1273 and 62,427 received the third, mRNA-1273 booster dose. The median date at the first dose was May 28, 2021, June 27, 2021 at the second dose, and December 22, 2021 at the third dose. The median time elapsed between the first and second doses was 28 days (IQR, 28-30 days). The median time elapsed between the second and booster doses was 228 days (IQR, 211-241 days).

As of January 9, 2022, 35,599 and 2,631 breakthrough infections were recorded among those who received either only two or three mRNA-1273 doses, respectively. Of the infections after the second dose, but before the booster dose, 17 progressed to severe, 5 to critical, and none to fatal COVID-19. Of the infections after the booster dose, 3 progressed to severe, but none to critical or fatal COVID-19.

Figure 1 shows the population selection process for the mRNA-1273-booster study. Table 1 shows the baseline characteristics of the full and matched cohorts. Median age in the matched cohorts was 40 years (IQR, 33-46). The matched cohorts were balanced on the matching factors.

### Effectiveness of mRNA-1273 booster against Omicron

The median time of follow-up was 14 days (IQR, 5-26 days) for the booster-dose and primary-series mRNA-1273 cohorts. A total of 1,463 infections, 317 symptomatic and 1,146 non-symptomatic at diagnosis, were recorded in the booster-dose cohort 8 days or more after receiving the booster dose (Figure 1). None of these infections progressed to severe, critical, or fatal COVID-19 as of end of this study. A total of 1,880 infections, 589 symptomatic and 1,291 non-symptomatic, were recorded in the primary-series cohort. None of these infections progressed to severe, critical, or fatal COVID-19.

Cumulative incidence of symptomatic infection was estimated at 1.9% (95% CI: 1.7-2.2%) for the booster-dose cohort and at 3.5% (95% CI: 3.2-3.9%) for the primary-series cohort, 35 days after the start of follow-up (Figure 2). Infection incidence was overwhelmingly predominated by Omicron. The median date of start of follow-up was December 23, 2021, after onset of the Omicron-wave exponential-growth phase on December 19, 2021.^10,36^

Symptomatic infection incidence rate was estimated at 32.8 (95% CI: 29.4-36.6) per 10,000 person-weeks in the booster-dose cohort and at 60.6 (95% CI: 55.9-65.7) per 10,000 person-weeks in the primary-series cohort (Table 2). The adjusted hazard ratio for symptomatic infection was estimated at 0.49 (95% CI: 0.43-0.57). Effectiveness of the mRNA-1273 booster dose relative to the primary series was estimated at 50.8% (95% CI: 43.4-57.3%).

mRNA-1273 vaccine effectiveness against any severe, critical, or fatal COVID-19 could not be calculated because there were no recorded cases in both the booster and primary-series cohorts as of end of this study (Figure 1).

### Additional analyses

For the BNT162b2-vaccine analysis, with the start of follow-up on the 15^th^ day after the booster dose, effectiveness of the booster dose against symptomatic (Omicron) infection, relative to the primary series, was estimated at 50.3% (95% CI: 47.5-53.0%) (Figure S1 and Table S2). The corresponding effectiveness for mRNA-1273 was estimated at 50.1% (95% CI: 41.4-57.6%) (Figure S1 and Table S2). Both effectiveness estimates were similar to the main analysis estimates.

Considering any documented (Omicron) infection, regardless of presence of symptoms or reason for PCR testing, as the primary outcome, effectiveness of the booster dose relative to the primary series was estimated at 38.2% (95% CI: 36.3-40.0%) for BNT162b2 and at 28.8% (95% CI: 23.6-33.6%) for mRNA1273.

During follow-up in the BNT162b2-vaccine analysis, 26.6% of individuals in the booster-dose cohort and 23.1% of individuals in the primary-series cohort had a PCR test done. During follow-up in the mRNA-1273-vaccine analysis, 21.3% of individuals in the booster-dose cohort and 14.6% of individuals in the primary-series cohort had a PCR test done. Those in the booster dose cohorts had higher testing rates due to higher pre-travel testing (Table S3). It appears that many individuals took the booster dose in preparation for planned travel during the year end holidays season.

Adjusting the effectiveness estimates for any documented (Omicron) infection by the ratio of testing rates across the booster and primary-series cohorts (Table S3), effectiveness of the booster dose relative to the primary series was estimated at 47.7% (95% CI: 46.0-49.3%) for BNT162b2 and at 54.0% (95% CI: 50.7-57.2%) for mRNA-1273.

## Discussion

BNT162b2 booster vaccination was associated with an 86% reduction in incidence of symptomatic infection for the Delta variant, and 50% reduction for the Omicron variant. For the mRNA-1273 booster, the reduction in incidence of Omicron was also similar at 51%. There were fewer cases of severe COVID-19 in the booster-dose cohorts than in the primary-series cohorts, consistent with strong protection against hospitalization and death for the booster, but cases of severe COVID-19 were rare in both booster-dose and primary-series cohorts. This affirms the durability of considerable vaccine protection against hospitalization and death several months after receiving the second dose.^1,6^ Although there was a large number of infections among those who received the booster, the number of severe COVID-19 cases was small and there were no cases of critical or fatal COVID-19.

These findings, in the context of a global expansion of Omicron and dwindling incidence of Delta, suggest that a longer term strategy for a global response is development and administration of a new generation of vaccines targeting a broad range of variants, or pan-coronavirus vaccines,^39,40^ to confront future SARS-CoV-2 variants, rather than continuing with a strategy of repeated booster vaccination employing existing vaccines. To illustrate how Omicron has eroded the effectiveness of current mRNA vaccines, the cumulative incidence of any documented infection in the vaccinated cohorts in Qatar was <1% after 6 months of follow-up during times in which incidence was predominated by Beta and Delta,^3^ but reached ten-fold higher in the present study to approximately 10% among recently boosted persons, after only 2-3 weeks of follow-up during the Omicron wave.

The above estimated effectiveness measures for Delta and Omicron are broadly consistent with growing evidence for effectiveness of mRNA vaccines against these variants in other countries,^26-29,41-52^ but in our study, effectiveness of boosters was compared to that of the primary series, and not to unvaccinated persons. Having said so, with the waning of vaccine protection against infection over time after second dose,^1,6^ the effectiveness among those boosted compared to those unvaccinated may not be much higher than that compared to those with only primary series. Effectiveness in this study was also assessed against specifically symptomatic infection, with results being also presented for any documented infection, symptomatic or asymptomatic.

This study has limitations. With the high and durable effectiveness of BNT162b2 and mRNA-1273 primary series against hospitalization and death,^1,6^ the lower severity of Omicron,^53^ the relatively young population of Qatar,^18,54^ and the time lag between infection and severe forms of COVID-19, there were too few confirmed severe, critical, and fatal COVID-19 cases at time of writing of this report to be able to precisely estimate effectiveness of the boosters against COVID-19 hospitalization and death. As an observational study, the vaccinated cohorts were neither blinded nor randomized, so potentially unmeasured or uncontrolled confounding cannot be excluded. While matching was done for sex, age, nationality, and calendar week of the second vaccine dose, it was not possible for other factors, such as comorbidities or occupation, as such data were not available to investigators. With the large Omicron wave in Qatar, use of rapid antigen testing was expanded to supplement PCR testing starting from January 5, 2022, but rapid antigen testing data were not available for analysis.

Matching was done to control for factors known to affect infection risk of exposure in Qatar,^18,21,30-32^ as well as variant exposure.^1,13,14,23,36-38^ Matching by age may have (partially) reduced the potential for bias due to co-morbidities. The number of individuals with severe chronic conditions is also small in Qatar’s young population.^18,54^ Matching by nationality may have (partially) controlled the differences in occupational risk, in consideration of the association between nationality and occupation in Qatar.^18,21,30-32^ The prescription for matching was recently shown to provide adequate control of the differences in the risk of exposure to the infection.^3^ Use of rapid antigen testing may not have differentially affected the investigated cohorts. A strength of this study is exclusion of those with a documented prior infection, as presence of prior infection may affect the protection afforded by the booster.^15^

In conclusion, mRNA boosters are associated with high effectiveness against Delta infection, but lower effectiveness against Omicron infection. There were fewer cases of severe COVID-19 in the booster-dose cohorts than in the primary-series cohorts, consistent with strong protection against hospitalization and death for the booster. Against future SARS-CoV-2 waves that may be driven by newly emerging variants, these findings suggest that a longer term strategy for a global response is development and administration of a new generation of vaccines targeting a broad range of variants, or pan-coronavirus vaccines, to confront future SARS-CoV-2 variants, rather than continuing with a strategy of repeated booster vaccination employing existing vaccines.

## Data Availability

The dataset of this study is a property of the Qatar Ministry of Public Health that was provided to the researchers through a restricted-access agreement that prevents sharing the dataset with a third party or publicly. Future access to this dataset can be considered through a direct application for data access to Her Excellency the Minister of Public Health (https://www.moph.gov.qa/english/Pages/default.aspx). Aggregate data are available within the manuscript and its Supplementary information.

## Acknowledgements

We acknowledge the many dedicated individuals at Hamad Medical Corporation, the Ministry of Public Health, the Primary Health Care Corporation, Qatar Biobank, Sidra Medicine, and Weill Cornell Medicine-Qatar for their diligent efforts and contributions to make this study possible. The authors are grateful for institutional salary support from the Biomedical Research Program and the Biostatistics, Epidemiology, and Biomathematics Research Core, both at Weill Cornell Medicine-Qatar, as well as for institutional salary support provided by the Ministry of Public Health, Hamad Medical Corporation, and Sidra Medicine. The authors are also grateful for the Qatar Genome Programme and Qatar University Biomedical Research Center for institutional support for the reagents needed for the viral genome sequencing. The funders of the study had no role in study design, data collection, data analysis, data interpretation, or writing of the article. Statements made herein are solely the responsibility of the authors.

## Author contributions

LJA conceived and co-designed the study, led the statistical analyses, and co-wrote the first draft of the article. HC co-designed the study, performed the statistical analyses, and co-wrote the first draft of the article. HY, HAK, and MS conducted viral genome sequencing. PT and MRH conducted the multiplex, RT-qPCR variant screening and viral genome sequencing. All authors contributed to data collection and acquisition, database development, discussion and interpretation of the results, and to the writing of the manuscript. All authors have read and approved the final manuscript.

## Supplementary Appendix

### Section 1. COVID-19 severity, criticality, and fatality classification

Severe Coronavirus Disease 2019 (COVID-19) disease was defined per the World health Organization (WHO) classification as a severe acute respiratory syndrome coronavirus 2 (SARS-CoV-2) infected person with “oxygen saturation of <90% on room air, and/or respiratory rate of >30 breaths/minute in adults and children >5 years old (or ≥60 breaths/minute in children <2 months old or ≥50 breaths/minute in children 2-11 months old or ≥40 breaths/minute in children 1–5 years old), and/or signs of severe respiratory distress (accessory muscle use and inability to complete full sentences, and, in children, very severe chest wall indrawing, grunting, central cyanosis, or presence of any other general danger signs)”.^1^ Detailed WHO criteria for classifying SARS-CoV-2 infection severity can be found in the WHO technical report.^1^

Critical COVID-19 disease was defined per WHO classification as a SARS-CoV-2 infected person with “acute respiratory distress syndrome, sepsis, septic shock, or other conditions that would normally require the provision of life sustaining therapies such as mechanical ventilation (invasive or non-invasive) or vasopressor therapy”.^1^ Detailed WHO criteria for classifying SARS-CoV-2 infection criticality can be found in the WHO technical report.^1^

COVID-19 death was defined per WHO classification as “a death resulting from a clinically compatible illness, in a probable or confirmed COVID-19 case, unless there is a clear alternative cause of death that cannot be related to COVID-19 disease (e.g. trauma). There should be no period of complete recovery from COVID-19 between illness and death. A death due to COVID-19 may not be attributed to another disease (e.g. cancer) and should be counted independently of preexisting conditions that are suspected of triggering a severe course of COVID-19”. Detailed WHO criteria for classifying COVID-19 death can be found in the WHO technical report.^2^

### Section 2. Laboratory methods

Nasopharyngeal and/or oropharyngeal swabs were collected for PCR testing and placed in Universal Transport Medium (UTM). Aliquots of UTM were: 1) extracted on a QIAsymphony platform (QIAGEN, USA), KingFisher Flex (Thermo Fisher Scientific, USA), MGISP-960 (MGI, China), or ExiPrep 96 Lite (Bioneer, South Korea) followed by testing with real-time reverse-transcription PCR (RT-qPCR) using TaqPath COVID-19 Combo Kits (Thermo Fisher Scientific, USA) on an ABI 7500 FAST (Thermo Fisher Scientific, USA); 2) tested directly on the Cepheid GeneXpert system using the Xpert Xpress SARS-CoV-2 (Cepheid, USA); or 3) loaded directly into a Roche cobas 6800 system and assayed with the cobas SARS-CoV-2 Test (Roche, Switzerland). The first assay targets the viral S, N, and ORF1ab gene regions. The second targets the viral N and E-gene regions, and the third targets the ORF1ab and E-gene regions.

All PCR testing was conducted at the Hamad Medical Corporation Central Laboratory or Sidra Medicine Laboratory, following standardized protocols.

**Table S1.** STROBE checklist for cohort studies..

|  | Item No | Recommendation | Main Text page |
| --- | --- | --- | --- |
| Title and abstract | 1 | (a) Indicate the study’s design with a commonly used term in the title or the abstract | Abstract |
|  |  | (b) Provide in the abstract an informative and balanced summary of what was done and what was found | Abstract |
| Introduction |  |  |  |
| Background/rationale | 2 | Explain the scientific background and rationale for the investigation being reported | Introduction |
| Objectives | 3 | State specific objectives, including any prespecified hypotheses | Introduction |
| Methods |  |  |  |
| Study design | 4 | Present key elements of study design early in the paper | Methods (‘Data sources and study design’, paragraph 2) |
| Setting | 5 | Describe the setting, locations, and relevant dates, including periods of recruitment, exposure, follow-up, and data collection | Methods (‘Data sources and study design’) & Figure 1 |
| Participants | 6 | (a) Give the eligibility criteria, and the sources and methods of selection of participants. Describe methods of follow-up<br>(b) For matched studies, give matching criteria and number of exposed and unexposed | Methods (‘Data sources and study design’, paragraphs 1-6) & Figure 1 |
| Variables | 7 | Clearly define all outcomes, exposures, predictors, potential confounders, and effect modifiers. Give diagnostic criteria, if applicable | Methods (‘Data sources and study design’, paragraphs 2-8) & Table 1 |
| Data sources/measurement | 8* | For each variable of interest, give sources of data and details of methods of assessment (measurement). Describe comparability of assessment methods if there is more than one group | Methods (‘Data sources and study design’ & ‘Laboratory methods’), Table 1 & Sections 1-2 in Appendix |
| Bias | 9 | Describe any efforts to address potential sources of bias | Methods (‘Data sources and study design’, paragraphs 4-5 & ‘Statistical analysis’, paragraph 4) |
| Study size | 10 | Explain how the study size was arrived at | Figure 1 |
| Quantitative variables | 11 | Explain how quantitative variables were handled in the analyses. If applicable, describe which groupings were chosen and why | Methods (‘Data sources and study design’, paragraphs 4 and 8) & Table 1 |
| Statistical methods | 12 | (a) Describe all statistical methods, including those used to control for confounding | Methods (‘Statistical analysis’) |
|  |  | (b) Describe any methods used to examine subgroups and interactions | NA |
|  |  | (c) Explain how missing data were addressed | NA, see Methods (‘Data sources and study design’, paragraph 1) |
|  |  | (d) If applicable, explain how loss to follow-up was addressed | NA, see Methods (‘Data sources and study design’, paragraph 1) |
|  |  | (e) Describe any sensitivity analyses | Methods (‘Data sources and study design’, paragraph 2 & ‘Statistical analysis’, paragraphs 3-4) |
| Results |  |  |  |
| Participants | 13* | (a) Report numbers of individuals at each stage of study—eg numbers potentially eligible, examined for eligibility, confirmed eligible, included in the study, completing follow-up, and analysed<br>(b) Give reasons for non-participation at each stage<br>(c) Consider use of a flow diagram | Results (‘Study population for BNT162b2 vaccine’ & ‘Study population for mRNA-1273 vaccine), Figure 1, & Table 1 |
| Descriptive data | 14 | (a) Give characteristics of study participants (eg demographic, clinical, social) and information on exposures and potential confounders | Results (‘Study population for BNT162b2 vaccine’ & ‘Study population for mRNA-1273 vaccine) & Table 1 |
|  |  | (b) Indicate number of participants with missing data for each variable of interest | NA, see Methods (‘Data sources and study design’, paragraph 1) |
|  |  | (c) Summarise follow-up time (eg, average and total amount) | Results (‘Study population for BNT162b2 vaccine’ & ‘Study population for mRNA-1273 vaccine), Figure 2, & Table 2 |
| Outcome data | 15 | Report numbers of outcome events or summary measures over time | Results (‘Effectiveness of BNT162b2 booster against Omicron’ & ‘Effectiveness of mRNA-1273 booster against Omicron). Figures 1-2, & Table 2 |
| Main results | 16 | (a) Give unadjusted estimates and, if applicable, confounder-adjusted estimates and their precision (eg, 95% confidence interval). Make clear which confounders were adjusted for and why they were included | Results ('Effectiveness of BNT162b2 booster against Omicron' & 'Effectiveness of mRNA-1273 booster against Omicron'), Figure 2, & Table 2 |
|  |  | (b) Report category boundaries when continuous variables were categorized | Table 1 |
|  |  | (c) If relevant, consider translating estimates of relative risk into absolute risk for a meaningful time period | NA |
| Other analyses | 17 | Report other analyses done—eg analyses of subgroups and interactions, and sensitivity analyses | Results ('Effectiveness of BNT162b2 booster against Delta' & 'Additional analyses'), Figure 3, Table 3, & Figure S1 & Tables S2-S3 in Appendix |
| Discussion |  |  |  |
| Key results | 18 | Summarise key results with reference to study objectives | Discussion, paragraphs 1-3 |
| Limitations | 19 | Discuss limitations of the study, taking into account sources of potential bias or imprecision. Discuss both direction and magnitude of any potential bias | Discussion, paragraphs 4-5 |
| Interpretation | 20 | Give a cautious overall interpretation of results considering objectives, limitations, multiplicity of analyses, results from similar studies, and other relevant evidence | Discussion, paragraph 6 |
| Generalisability | 21 | Discuss the generalisability (external validity) of the study results | Discussion, paragraphs 4-5 |
| Other information |  |  |  |
| Funding | 22 | Give the source of funding and the role of the funders for the present study and, if applicable, for the original study on which the present article is based | Acknowledgements |

**Figure S1.**
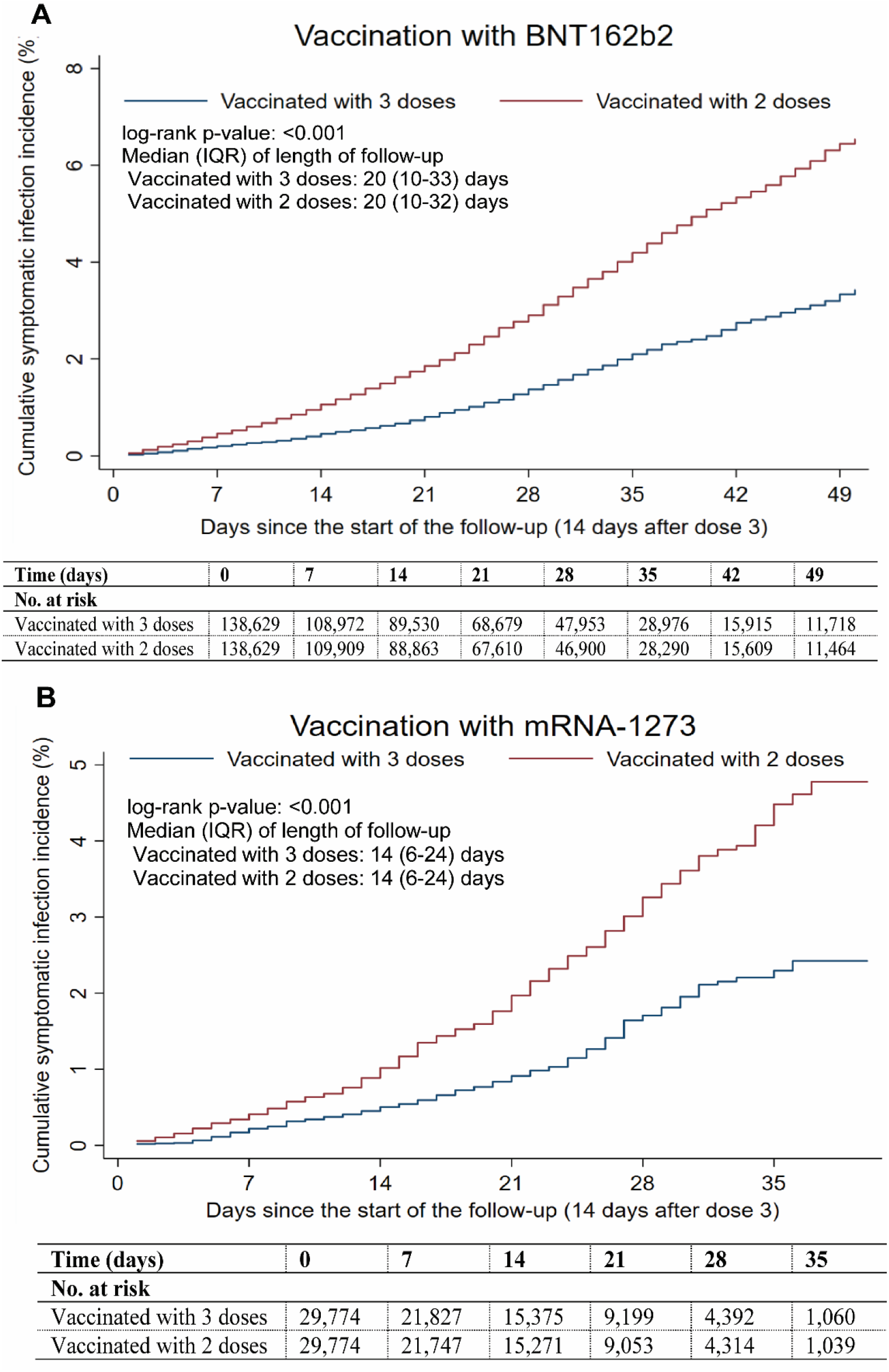
Cumulative incidence of symptomatic infection in the matched booster-dose and primary-series cohorts for each of the BNT162b2 (A) and mRNA-1273 (B) vaccines, with start of follow-up on the 15^th^ day after booster dose.

**Table S2.** Effectiveness of BNT162b2 and mRNA-1273 booster vaccination, with start of follow-up on the 15^th^ day after booster dose..

| Epidemiological measure | BNT162b2 |  | mRNA-1273 |  |
| --- | --- | --- | --- | --- |
|  | Estimate (95% CI) | Effectiveness in %<br>(95% CI) | Estimate (95% CI) | Effectiveness in %<br>(95% CI) |
| Total follow-up time—booster-dose cohort (person-weeks) | 465,476 | -- | 65,523 | -- |
| Total follow-up time—primary-series cohort (person-weeks) | 460,012 | -- | 65,023 | -- |
| Incidence rate for symptomatic infection—booster-dose cohort (per 10,000 person-weeks) | 39.9 (38.1-41.8) | -- | 35.7 (31.4-40.6) | -- |
| Incidence rate for symptomatic infection—primary-series cohort (per 10,000 person-weeks) | 78.5 (76.0-81.1) | -- | 67.8 (61.8-74.5) | -- |
| Unadjusted hazard ratio for symptomatic infection | 0.50 (0.47-0.53) | 50.0 (47.1-52.7) | 0.50 (0.43-0.59) | 50.0 (41.2-57.4) |
| Adjusted hazard ratio for symptomatic infection* | 0.50 (0.47-0.53) | 50.3 (47.5-53.0) | 0.50 (0.42-0.59) | 50.1 (41.4-57.6) |

Abbreviations: CI, confidence interval.
\*Cox regression analysis adjusted for sex, 10 age-groups, 10 nationality groups, and calendar week of second vaccine dose.

**Table S3.** Distribution of the reason for PCR testing during follow-up for A) all PCR tests during follow-up and B) identified infections, among the matched booster-dose and primary-series cohorts for the BNT162b2 and mRNA-1273 vaccines.

|  | BNT162b2 |  |  |  |  |  | mRNA-1273 |  |  |  |  |  |
| --- | --- | --- | --- | --- | --- | --- | --- | --- | --- | --- | --- | --- |
|  | A) All PCR tests |  |  | B) PCR-confirmed infections |  |  | A) All PCR tests |  |  | B) PCR-confirmed infections |  |  |
|  | Individuals who received Dose 3* | Individuals who did not receive Dose 3* | SMD† | Individuals who received Dose 3 | Individuals who did not receive Dose 3 | SMD† | Individuals who received Dose 3* | Individuals who did not receive Dose 3* | SMD† | Individuals who received Dose 3 | Individuals who did not receive Dose 3 | SMD† |
| Number of tests | N=65,589 | N=55,369 |  | N=7,210 | N=10,994 |  | N=10,742 | N=6,962 |  | N=1,463 | N=1,880 |  |
| **Reason for PCR testing** |  |  |  |  |  |  |  |  |  |  |  |  |
| Clinical suspicion | 5,677 (8.7) | 7,169 (13.0) | 0.28 | 2,035 (28.2) | 3,858 (35.1) | 0.20 | 824 (7.7) | 926 (13.3) | 0.43 | 317 (21.7) | 589 (31.3) | 0.30 |
| Contact tracing | 2,930 (4.5) | 2,791 (5.0) |  | 867 (12.0) | 1,080 (9.8) |  | 359 (3.3) | 409 (5.9) |  | 124 (8.5) | 196 (10.4) |  |
| Survey | 6,847 (10.4) | 7,377 (13.3) |  | 951 (13.2) | 1,570 (14.3) |  | 821 (7.6) | 856 (12.3) |  | 180 (12.3) | 233 (12.4) |  |
| Individual request | 2,268 (3.5) | 2,005 (3.6) |  | 421 (5.8) | 644 (5.9) |  | 509 (4.7) | 322 (4.6) |  | 100 (6.8) | 125 (6.7) |  |
| Pre-travel | 34,361 (52.4) | 21,683 (39.2) |  | 2,288 (31.7) | 2,811 (25.6) |  | 6,462 (60.2) | 2,775 (39.9) |  | 667 (45.6) | 619 (32.9) |  |
| Port of entry | 12,349 (18.8) | 12,893 (23.3) |  | 455 (6.3) | 680 (6.2) |  | 1,674 (15.6) | 1,554 (22.3) |  | 60 (4.1) | 82 (4.4) |  |
| Healthcare routine testing | 959 (1.5) | 1,335 (2.4) |  | 150 (2.1) | 316 (2.9) |  | 86 (0.8) | 103 (1.5) |  | 13 (0.9) | 31 (1.7) |  |
| Other | 198 (0.3) | 116 (0.2) |  | 43 (0.6) | 35 (0.3) |  | 7 (0.1) | 17 (0.2) |  | 2 (0.1) | 5 (0.3) |  |

Abbreviations: SMD, standardized mean difference.
\*Cohorts were matched one-to-one by sex, 10-year age group, nationality, and calendar week of second vaccine dose.
†SMD is the difference in the mean of a covariate between groups divided by the pooled SD.

